# Development and calibration of a simple mortality risk score for hospitalized COVID-19 adults

**DOI:** 10.1101/2020.08.31.20185363

**Authors:** Edwin Yoo, Bethany Percha, Max Tomlinson, Victor Razuk, Stephanie Pan, Madeleine Basist, Pranai Tandon, Jing Gennie Wang, Cynthia Gao, Sonali Bose, Umesh Gidwani

## Abstract

**Objectives:** Mortality risk scores, such as SOFA, qSOFA, and CURB-65, are quick, effective tools for communicating a patient’s prognosis and guiding therapeutic decisions. Most use simple calculations that can be performed by hand. While several COVID-19 specific risk scores exist, they lack the ease of use of these simpler scores. The objectives of this study were (1) to design, validate, and calibrate a simple, easy-to-use mortality risk score for COVID-19 patients and (2) to recalibrate SOFA, qSOFA, and CURB-65 in a hospitalized COVID-19 population.

**Design:** Retrospective cohort study incorporating demographic, clinical, laboratory, and admissions data from electronic health records.

**Setting:** Multi-hospital health system in New York City. Five hospitals were included: one quaternary care facility, one tertiary care facility, and three community hospitals.

**Participants:** Patients (n=4840) with laboratory-confirmed SARS-CoV2 infection who were admitted between March 1 and April 28, 2020.

**Main outcome measures:** Gray’s K-sample test for the cumulative incidence of a competing risk was used to assess and rank 48 different variables’ associations with mortality. Candidate variables were added to the composite score using DeLong’s test to evaluate their effect on predictive performance (AUC) of in-hospital mortality. Final AUCs for the new score, SOFA, qSOFA, and CURB-65 were assessed on an independent test set.

**Results:** Of 48 variables investigated, 36 (75%) displayed significant (p<0.05 by Gray’s test) associations with mortality. The variables selected for the final score were (1) oxygen support level, (2) troponin, (3) blood urea nitrogen, (4) lymphocyte percentage, (5) Glasgow Coma Score, and (6) age. The new score, COBALT, outperforms SOFA, qSOFA, and CURB-65 at predicting mortality in this COVID-19 population: AUCs for initial, maximum, and mean COBALT scores were 0.81, 0.91, and 0.92, compared to 0.77, 0.87, and 0.87 for SOFA. We provide COVID-19 specific mortality estimates at all score levels for COBALT, SOFA, qSOFA, and CURB-65.

**Conclusions:** The COBALT score provides a simple way to estimate mortality risk in hospitalized COVID-19 patients with superior performance to SOFA and other scores currently in widespread use. Evaluation of SOFA, qSOFA, and CURB-65 in this population highlights the importance of recalibrating mortality risk scores when they are used under novel conditions, such as the COVID-19 pandemic. This study’s approach to score design could also be applied in other contexts to create simple, practical and high-performing mortality risk scores.

**Trial registration:** NA

**Funding source:** The authors declare that there was no external funding provided.

**Summary box:** *What is already known on this topic:*

- Mortality risk scores are widely used in clinical settings to facilitate communication with patients and families, guide goals of care discussions, and optimize resource allocation.
- Although popular mortality risk scores like SOFA, qSOFA, and CURB-65 are routinely used in COVID-19 populations, they were originally calibrated in different contexts and their true performance among hospitalized COVID-19 patients is unknown.
- Several dedicated COVID-19 mortality risk scores have been created during the 2019-2020 pandemic, but all use complicated formulae or machine learning algorithms and are difficult or impossible to calculate by hand, limiting their applicability at the bedside.

*What this study adds:*

- We describe a data-driven, simple, and hand-calculable COVID-specific mortality risk score (COBALT) that has superior performance to SOFA, qSOFA, and CURB-65 in a hospitalized COVID-19 patient population.
- We provide COVID-specific mortality estimates for SOFA, qSOFA, and CURB-65 using data from 4840 patients in a large and diverse New York City multihospital health system.

## Introduction

Mortality risk scores are widely used in clinical settings to facilitate communication with patients and families, guide goals of care discussions, and improve triage under resource constraints. When used appropriately, they provide an objective, realistic assessment of a patient’s prognosis [1]. Such objectivity is especially important under conditions of uncertainty, such as the 2019 coronavirus (COVID-19) pandemic, which has claimed the lives of over 142,000 people in the United States and over 618,000 worldwide [2]. When family members cannot be with their loved ones directly, or when essential resources such as ICU beds and ventilators are limited, physicians’ ability to transparently communicate mortality risk becomes increasingly critical [3].

There are two approaches to the use of mortality risk scores in a novel context, such as COVID-19. One is to repurpose scores that are already in widespread use in similar patient populations, such as SOFA [4], qSOFA [5], and CURB-65 [6]. The other is to develop a score de novo, either through expert assessment of risk factors for COVID-19 mortality or through data-driven methods. Many widely used scores, like SOFA, qSOFA, and CURB-65, have been extensively validated [7-10] and are easy to calculate.

However, further calibration in COVID-19 populations is necessary to ensure that the estimates of mortality risk at each score level are accurate. The second approach, designing a new score, has been much more widely pursued in COVID-19. Unfortunately, many of these new scores have been found to be biased, poorly evaluated, or otherwise unsuitable [11]. In addition, even well-designed and high performing scores can suffer from limited practical utility. For example, to our knowledge, all existing scores designed specifically for COVID-19 involve complicated formulae or machine learning algorithms and are deployed using online calculators [12-16,30]. This limits their applicability, especially in time- and resource-constrained settings.

Given these considerations, we sought to improve COVID-19 mortality risk prediction in two ways. First, we generated COVID-19 specific mortality estimates for three existing mortality risk scores - SOFA, qSOFA, and CURB-65 - using the electronic health records (EHRs) of 4840 COVID-19 patients who were admitted to the Mount Sinai Health System in New York City during March and April 2020. Second, we developed a method for creating simple, additive mortality risk scores from EHR data. We then applied this method to create a new six-variable score (“COBALT”) that predicts mortality among hospitalized COVID-19 patients with greater accuracy than SOFA, qSOFA, and CURB-65 while maintaining their ease of use. Both the COBALT score and the recalibrated mortality estimates for the other three scores should substantially improve clinicians’ ability to understand and communicate mortality risk in COVID-19.

## Methods

### Study population and dataset

This study was approved by the Mount Sinai Institutional Review Board (IRB-20-03613). A graphical summary of the dataset preparation is shown in **Figure 1**. The study population consisted of 4840 patients between the ages of 18 and 99 who were admitted to one of five hospitals within the Mount Sinai Health System between March 1 and April 28, 2020 and who had a positive PCR test for SARS-CoV2 during or prior to their admissions. Patients’ admission histories, demographics, laboratory tests, oxygen device use, vital signs, comorbidities, and medications were extracted from electronic databases (Epic Caboodle and Clarity; Epic Systems Corporation). If a patient was transferred from one Mount Sinai hospital to another, we included both admissions. The final outcome (death or discharge) was assessed using the recorded discharge disposition at the end of the patient’s admission. The study follow-up period ended on May 31, 2020 and patients still admitted at this time were considered censored.

**Figure 1:**
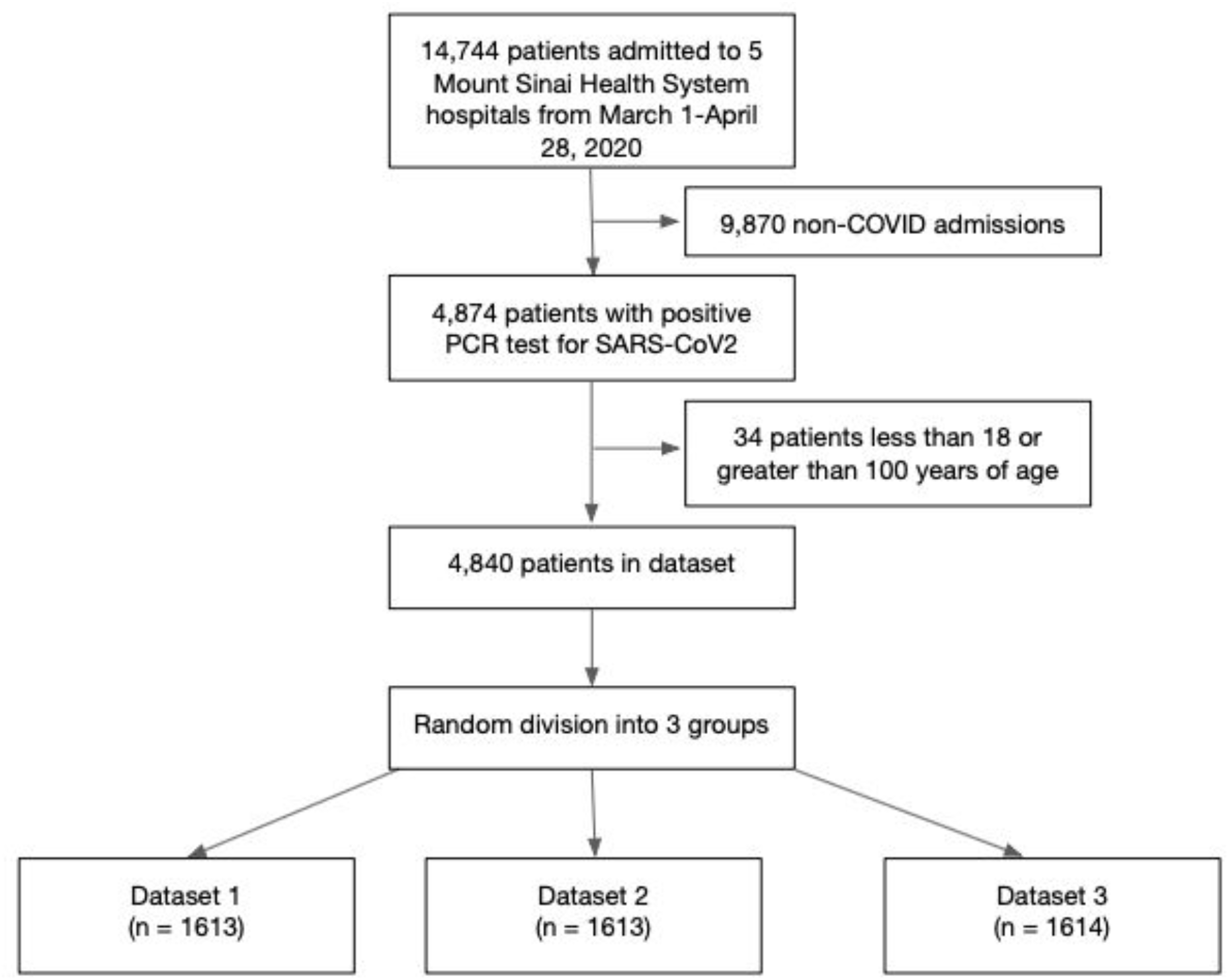
Flow chart of study design.

Each patient’s clinical history was represented as a series of time intervals ranging from the time of admission to the time of discharge, death, or censoring. The creation of a new interval was triggered by any change in a patient’s laboratory values, vital signs, oxygen support devices, or oxygen settings (e.g. FiO_2_) or any other time-varying variable. Non-time-varying information about each patient, including age at admission, sex, admission site, smoking status, self-reported race and ethnicity, and comorbidities, was also included. **Table 1** contains a complete list of the variables considered in this study.

**Table 1:** Characteristics of study population. Reported laboratory values and vital signs are those collected closest to the patient’s time of admission.

| Characteristic | Value<br>(n = 4840) |
| --- | --- |
| **Age (years) – median (IQR)** | 66.4 (54.9, 77.8) |
| **Sex – No. (%)** |  |
| - Male | 2733 (56.5) |
| - Female | 2107 (43.5) |
| **Race – No. (%)** |  |
| - White | 1190 (24.6) |
| - Black | 1342 (27.7) |
| - Asian/Pacific Islander | 238 (4.9) |
| - Other | 1919 (39.7) |
| - Unknown | 151 (3.1) |
| **Ethnicity – No. (%)** |  |
| - Hispanic | 1345 (27.8) |
| - Non-Hispanic | 2861 (59.1) |
| - Unknown | 634 (13.1) |
| **Hospital of Admission** |  |
| - Hospital 1 (quaternary care facility) | 1762 (36.4) |
| - Hospital 2 (community hospital) | 821 (17.0) |
| - Hospital 3 (community hospital) | 800 (16.5) |
| - Hospital 4 (tertiary care hospital) | 561 (11.6) |
| - Hospital 5 (community hospital) | 896 (18.5) |
| **Insurance Status** |  |
| - Medicaid | 751 (15.5) |
| - Medicare | 1948 (40.2) |
| - Private | 1089 (22.5) |
| - Self-Pay | 109 (2.3) |
| - Other | 392 (8.1) |
| - Unknown | 551 (11.4) |
| **Comorbidities – No. (%)** |  |
| - Hypertension | 2232 (46.1) |
| - Cardiovascular diseases<br>- Coronary artery disease<br>- Heart failure<br>- Chronic lung diseases<br>- Asthma<br>- COPD/Emphysema<br>- Diabetes mellitus type 2<br>- Chronic kidney disease<br>- Bleeding or clotting disorder<br>- Cerebrovascular disease<br>- Cancer<br>- Liver disease<br>- Pregnancy<br>- Prior transplant<br>- Human immunodeficiency virus (HIV) | 2131 (44.0)<br>821 (17.0)<br>704 (14.5)<br>2107 (43.5)<br>419 (8.7)<br>315 (6.5)<br>1513 (31.3)<br>839 (17.3)<br>706 (14.6)<br>532 (11.0)<br>510 (10.5)<br>239 (4.9)<br>156 (3.2)<br>129 (2.7)<br>99 (2.0) |
| **Smoking status – No. (%)**<br>- Current<br>- Former<br>- Never<br>- Unknown | 214 (4.4)<br>1035 (21.4)<br>2580 (53.3)<br>1011 (20.9) |
| **Baseline vitals – Median (IQR; number missing)**<br>- Body mass index (BMI)<br>- Diastolic blood pressure (mmHg)<br>- Systolic blood pressure (mmHg)<br>- Glasgow coma score<br>- Mean arterial pressure (MAP)<br>- Oxygen saturation (%)<br>- Heart rate (bpm)<br>- Respiratory rate<br>- Temperature (°F) | 28.0 (24.6-32.7; 927)<br>73.0 (65.0-82.0; 4)<br>128.0 (114.0-144.0; 4)<br>15.0 (12.0-15.0; 1932)<br>90.0 (81.0-100.0; 423)<br>95.0 (92.0-98.0; 4)<br>95 (82-109; 3)<br>20 (18-22; 6)<br>98.6 (98.0-99.9; 5) |
| **Baseline laboratory values – Median (IQR; number missing)**<br>- Albumin (g/dL)<br>- Bilirubin (mg/dL)<br>- Blood urea nitrogen (mg/dL) | 3.2 (2.8-3.5; 166)<br>0.6 (0.4-0.8; 166)<br>19 (12-35; 79) |
| - Creatinine (mg/dL) | 1.04 (0.80-1.64; 78) |
| - C reactive protein (mg/L) | 116.7 (56.1-203.4; 671) |
| - D-Dimer (µg/mL) | 1.71 (0.91-3.53; 865) |
| - Estimated glomerular filtration rate (EGFR) | 60 (36-60; 78) |
| - Ferritin (ng/mL) | 750 (340-1800; 723) |
| - Hemoglobin (g/dL) | 13.2 (11.7-14.5; 32) |
| - Interleukin-6 (IL6; pg/mL) | 70.9 (34.0-144.2; 2222) |
| - Lactic acid dehydrogenase (LDH; U/L) | 427 (319-584; 852) |
| - Lymphocyte percentage (%) | 12.1 (7.8-18.6; 74) |
| - PaO2 (arterial; mmHg) | 87 (64-131; 3527) |
| - Platelets (x10<sup>3</sup>/µL) | 211 (160-280; 36) |
| - Procalcitonin (ng/mL) | 0.19 (0.08-0.66; 980) |
| - Troponin (ng/mL) | 0.020 (0.010-0.065; 537) |
| - White blood cell count (x10<sup>3</sup>/µL) | 7.6 (5.6-10.6; 32) |
| **Baseline risk scores – Median (IQR)** |  |
| - SOFA | 1 (0-3) |
| - qSOFA | 0 (0-1) |
| - CURB-65 | 1 (1-2) |
| **Baseline oxygen requirement – No. (%)** |  |
| - Room air | 2029 (41.9) |
| - Low flow / non-rebreather | 2248 (46.5) |
| - High flow O2 | 74 (1.5) |
| - Non-invasive ventilation (NIV)<br>(e.g. CPAP/BiPAP) | 200 (4.1) |
| - Invasive mechanical ventilation (IMV) | 228 (4.7) |
| - NA / No record | 61 (1.3) |

The detailed nature of our time interval data allowed us to calculate the values of the SOFA [4], qSOFA [5], and CURB-65 [6] scores throughout each patient’s entire admission. For SOFA, oxygen support level (as estimated by delivery device), FiO_2_, mean arterial pressure, and the Glasgow Coma Scale (GCS) were obtained from nursing flowsheet data. Platelet count, bilirubin, creatinine, and PaO_2_ values came from laboratory result documentation. Vasopressor administration information came from the medication administration record (MAR); a patient was considered to be receiving vasopressor support from the documented time of administration until the order expired or was discontinued. Urine output was documented in nursing flowsheets as individual events; urine output over the last 24 hours was calculated by summing any documented urine output volumes within 24 hours of the start of the current interval. Additional information required for the qSOFA and CURB-65 scores included respiratory rate and blood pressure (systolic and diastolic), both of which were obtained from flowsheet documentation. CURB-65 includes a parameter for “confusion”; because this was not separately documented outside of unstructured clinical notes, we took it to mean a GCS score of less than 15.

We randomly divided the patients into three groups. Dataset 1 (n=1613) was used to quantify each variable’s association with mortality and choose preliminary candidates for the COBALT score. Dataset 2 (n=1613) was used to evaluate combinations of variables and select the final set of variables for the score. Dataset 3 (n=1614) was used to evaluate the final performance of the score and compare it to SOFA, qSOFA, and CURB-65. All three datasets were used to calculate mortality risk estimates for the four scores. Supplementary Table S1 confirms that the distributions of study variables were consistent across the three datasets.

### Selection of candidate variables and weights for the COBALT score

To identify promising variables for a COVID-specific mortality risk score, we sought those for which the associated mortality changed significantly across the observed values of the variable. For categorical variables this meant significant variation in mortality across categories, while for continuous variables it meant significant variation across the 0-25%, 25-50%, 50-75%, and 75-100% percentile ranges.

We created 100 random variants of Dataset 1 by first creating a bootstrap sample of the patients and then sampling a single time point uniformly from within each patient’s record. The values of 48 variables (**Tables 1-2**) were obtained at the sampled times, and the cumulative incidences of mortality across the different levels of each variable (starting at the sampled times) were compared using Gray’s K-sample tests for the cumulative incidence of a competing risk [17-18], stratifying by site of admission and treating hospital discharge to home and hospital discharge to a facility as competing risks. Gray’s tests provide a test statistic and p-value that quantify the difference in cumulative incidence among groups; the 48 variables were ranked according to this p-value. Only those sampled patients for whom the variable in question had been measured were included in each test; we did not attempt to impute values or otherwise interpret missing values.

**Table 2:** Tests of variables’ association with mortality and selection of COBALT candidate variables. The cumulative incidences of mortality among the 0-25%, 25-50%, 50-75%, and 75-100% percentiles of each variable were compared. The variables selected for further evaluation are highlighted in gray. Availability is summarized as follows: 1 = no blood required; 2 = blood required, routine lab; 3 = blood required, specialized lab.

|  | Parameter | Percentiles<br>(25%, 50%, 75%) | Test<br>Statistic<br>(Median) | P-value<br>(Median) | Availability |
| --- | --- | --- | --- | --- | --- |
| 1 | Oxygen support level (0-4) | Categorical | 392.7 | <0.0001 | 1 |
| 2 | Blood urea nitrogen (mg/dL) | 12.0, 20.0, 41.0 | 326.7 | <0.0001 | 2 |
| 3 | Lymphocyte percentage (%) | 7.8, 13.2, 20.7 | 266.9 | <0.0001 | 2 |
| 4 | Troponin (ng/mL) | 0.010, 0.020, 0.073 | 252.7 | <0.0001 | 2 |
| 5 | Respiratory rate | 18, 18, 20 | 250.7 | <0.0001 | 1 |
| 6 | Glasgow Coma Scale (GCS) | 9, 14, 15 | 227.4 | <0.0001 | 1 |
| 7 | Creatinine (mg/dL) | 0.71, 0.96, 1.63 | 218.8 | <0.0001 | 2 |
| 8 | EGFR | 36.0, 60.0, 60.0 | 212.3 | <0.0001 | 2 |
| 9 | White blood cell count (x1000/μL) | 5.6, 8.0, 11.3 | 199.2 | <0.0001 | 2 |
| 10 | Vasopressor administration | Binary | 153.3 | <0.0001 | 1 |
| 11 | Procalcitonin (ng/mL) | 0.07, 0.19, 0.77 | 152.6 | <0.0001 | 3 |
| 12 | CRP (mg/L) | 42.1, 94.8, 182.5 | 152.5 | <0.0001 | 3 |
| 13 | Age (years) | 55.2, 66.0, 78.2 | 147.0 | <0.0001 | 1 |
| 14 | LDH (U/L) | 316.0, 414.5, 566.0 | 144.0 | <0.0001 | 3 |
| 15 | Albumin (g/dL) | 2.4, 2.8, 3.2 | 122.2 | <0.0001 | 3 |
| 16 | D-Dimer (μg/mL) | 0.90, 1.85, 3.96 | 118.3 | <0.0001 | 3 |
| 17 | Interleukin-6 (pg/mL) | 30.1, 65.6, 135.3 | 82.7 | <0.0001 | 3 |
| 18 | Cardiovascular disease | Binary | 78.6 | <0.0001 | 1 |
| 19 | Heart rate (bpm) | 73, 84, 96 | 77.0 | <0.0001 | 1 |
| 20 | Diastolic blood pressure (mmHg) | 62, 70, 78 | 56.9 | <0.0001 | 1 |
| 21 | Ferritin (ng/mL) | 367.8, 782.0, 1792.6 | 49.2 | <0.0001 | 3 |
| 22 | Oxygen saturation (%) | 94, 96, 98 | 49.2 | <0.0001 | 1 |
| 23 | Systolic blood pressure (mmHg) | 110, 123, 137 | 31.9 | <0.0001 | 1 |
| 24 | Platelets (x10<sup>3</sup>/μL) | 170.8, 235.0, 325.0 | 31.6 | <0.0001 | 2 |
| 25 | PaO<sub>2</sub>/FiO<sub>2</sub> | 85.8, 145.7, 245.0 | 29.1 | <0.0001 | 3 |
| 26 | Coronary artery disease | Binary | 24.7 | <0.0001 | 1 |
| 27 | Hemoglobin (g/dL) | 10.4, 12.2, 13.6 | 23.8 | <0.0001 | 2 |
| 28 | Temperature (°F) | 97.5, 98.2, 99.0 | 22.5 | <0.0001 | 1 |
| 29 | Bilirubin (mg/dL) | 0.4, 0.6, 0.8 | 20.0 | 0.0002 | 2 |
| 30 | Hypertension | Binary | 13.3 | 0.0003 | 1 |
| 31 | Heart failure | Binary | 12.5 | 0.0004 | 1 |
| 32 | Chronic kidney disease | Binary | 6.4 | 0.012 | 1 |
| 33 | History of smoking | Binary | 6.0 | 0.014 | 1 |
| 34 | Diabetes mellitus II | Binary | 4.8 | 0.028 | 1 |
| 35 | Cancer/malignancy | Binary | 4.7 | 0.031 | 1 |
| 36 | Hispanic | Binary | 4.6 | 0.033 | 1 |
| 37 | Cerebrovascular disease | Binary | 3.0 | 0.083 | 1 |
| 38 | Prior transplant recipient | Binary | 3.0 | 0.086 | 1 |
| 39 | Black race | Binary | 2.7 | 0.10 | 1 |
| 40 | Emphysema/COPD | Binary | 1.6 | 0.21 | 1 |
| 41 | Chronic lung disease | Binary | 1.1 | 0.29 | 1 |
| 42 | Liver disease | Binary | 0.9 | 0.34 | 1 |
| 43 | Sex (Male/Female) | Binary | 0.7 | 0.40 | 1 |
| 44 | HIV | Binary | 0.7 | 0.40 | 1 |
| 45 | Body mass index | 24.6, 28.0, 32.6 | 2.9 | 0.41 | 1 |
| 46 | History of bleeding/clotting disorder | Binary | 0.6 | 0.43 | 1 |
| 47 | Current smoker | Binary | 0.6 | 0.44 | 1 |
| 48 | Asthma | Binary | 0.6 | 0.44 | 1 |

Abbreviations: *COPD* chronic obstructive pulmonary disease, *CRP* C-reactive protein, *EGFR* estimated glomerular filtration rate

The median cumulative incidences of mortality across quartiles/categories of each variable at 14 days after the time of measurement were used to determine the score weights (the values added to the score for each variable). The value for each quartile/category was multiplied by 10 and rounded to the nearest integer; the weight for the lowest mortality category was set to zero.

### Creation of the COBALT composite score

The top 20 variables identified by Gray’s tests were selected as potential score candidates. Using Dataset 2 and the score breaks and weights from Dataset 1, variables were added to the score one at a time. The associations between (a) value of the score at admission, (b) maximum value of the score during admission, and (c) mean value of the score during admission and final outcome (death = 1, other outcomes = 0) were assessed using the area under the receiver operating characteristic curve (AUC). At each stage, the remaining variable whose addition led to the greatest increase in AUC for the score measured at admission was identified. It was added to the composite score only if (a) DeLong’s test revealed a statistically significant (p<0.01) increase in AUC, (b) the absolute value of the AUC increased by more than 0.005, and (c) DeLong’s test did not reveal a statistically significant decline in AUC for the maximum or mean scores.

### Evaluation of score performance and creation of mortality tables

The same three AUC calculations used in the score selection process were applied to Dataset 3 and used to assess the performance of the COBALT score relative to SOFA, qSOFA, and CURB-65. Mortality rates across the ranges of each of the four scores (COBALT, SOFA, qSOFA, CURB-65) were calculated per hospital and for all patients using the combined dataset (Datasets 1-3) who had achieved an outcome at the end of the study period; 4751 out of 4840 patients, or 98.2%, were included.

### Software and libraries

All preprocessing of the raw electronic health record data was performed in Python (version 3.7.7). All statistical analysis was done in R (version 3.6.3). The Gray tests were performed in R using the cmprsk package [19].

## Results

### Characteristics of the study population and dataset

**Table 1** describes the baseline characteristics of all 4840 patients in our study population. The population was 56.5% male and 43.5% female with a median (IQR) age at admission of 66.4 (54.9-77.8) years. Patients self-reported race and ethnicity separately; 24.6% identified as white, 27.7% as Black, 4.9% as Asian/Pacific Islander, and 39.7% as “other”. Most of the patients listing their race as “other” reported Hispanic ethnicity; the study population was 27.8% Hispanic overall. There was good representation across the five Mount Sinai hospitals, which are situated in different neighborhoods across Manhattan, Queens, and Brooklyn. The most common comorbidities were hypertension (46.1%), followed by cardiovascular disease (e.g. coronary artery disease, heart failure; 44.0%), chronic lung diseases (e.g. asthma, chronic obstructive lung disease, emphysema; 43.5%), diabetes mellitus type 2 (31.3%), and chronic kidney disease (17.3%). Most patients (53.3%) had never smoked, and only 4.4% were current smokers. The majority did not require substantial oxygen support on admission: 41.9% were admitted on room air and 46.5% on low-flow oxygen (nasal cannula or non-rebreather mask).

### Parameter selection for the COBALT risk score

We considered 48 variables (labs, demographics, comorbidities, vital signs, oxygen support level, vasopressors; see **Table 2**) for inclusion in the risk score. Of these, 36 (75%) showed statistically significant (p<0.05) differences in the cumulative incidence of mortality across quartiles/categories. We chose the top 20 of these for further exploration. We later removed creatinine and EGFR from consideration because BUN, another measure of kidney function, showed a stronger association with mortality. We likewise removed respiratory rate because it was correlated with ventilation status, which was already represented by oxygen support level. Because so many variables were observed to impact mortality, it is likely that multiple combinations of variables could be used to construct useful scores and indeed, different combinations of variables often produced similar AUC values on Dataset 2 (data not shown). We therefore focused our attention on those variables that were most readily available: those that required no blood draw or were routine labs.

The final six variables selected for the COBALT score were Glasgow Coma Scale (“Coma” = “C”), oxygen support level (“Oxygen” = “O”), BUN (“B”), Age (“A”), lymphocyte percentage (“L”), and troponin (“T”). The COBALT score is similar in structure to SOFA and can be calculated using a simple additive formula as shown in **Table 3**.

**Table 3:** Calculation of the COBALT score. An example calculation is shown for a [fictitious] 65-year-old patient who is on high flow oxygen with BUN of 18.0 mg/dL, troponin 0.020 ng/mL, lymphocyte percentage 14%, no cognitive impairment (GCS=15).

| Parameter | Units | Criterion | Add | Example Calculation |
| --- | --- | --- | --- | --- |
| Oxygen delivery method |  | Room air<br>Nasal cannula / non-rebreather mask<br>High-flow nasal cannula<br>CPAP/BiPAP<br>Invasive mechanical ventilation (IMV) | 0<br>2<br>4<br>7<br>7 | 4 |
| Blood Urea Nitrogen (BUN) | mg/dL | ≤12.0 or Unknown<br>12.1-20.0<br>20.1-40.0<br>>40.0 | 0<br>1<br>3<br>6 | 1 |
| Troponin | ng/mL | ≤0.010 or Unknown<br>0.011-0.020<br>0.021-0.073<br>>0.073 | 0<br>2<br>3<br>6 | 2 |
| Lymphocyte percentage | % | >20.1 or Unknown<br>13.3-20.1<br>7.9-13.2<br>≤7.8 | 0<br>1<br>3<br>5 | 1 |
| Glasgow Coma Scale (GCS) |  | 15 or Unknown<br>14<br>7-13<br>≤6 | 0<br>1<br>3<br>7 | 0 |
| Age | years | <55 or Unknown<br>55-64<br>65-79<br>>80 | 0<br>2<br>3<br>5 | 3 |
|  |  |  | Range:<br>0-36 | TOTAL = 11 |

### Performance of SOFA, qSOFA, CURB-65, and COBALT scores

A comparison of COBALT to SOFA, qSOFA, and CURB-65 is shown in **Figure 2**. The COBALT score outperformed all three comparator scores regardless of whether the score used was the score at admission (AUCs: 0.81 for COBALT, 0.77 for SOFA, 0.75 for CURB-65, 0.69 for qSOFA), the maximum score over the patient’s admission (AUCs: 0.91 for COBALT, 0.87 for SOFA, 0.85 for CURB-65, 0.82 for qSOFA), or the mean score throughout the admission (AUCs: 0.91 for COBALT, 0.87 for SOFA, 0.87 for CURB-65, 0.85 for qSOFA). The COBALT score significantly outperformed SOFA (p<0.001 by DeLong’s test for admission, maximum, and mean scores).

**Figure 2:**
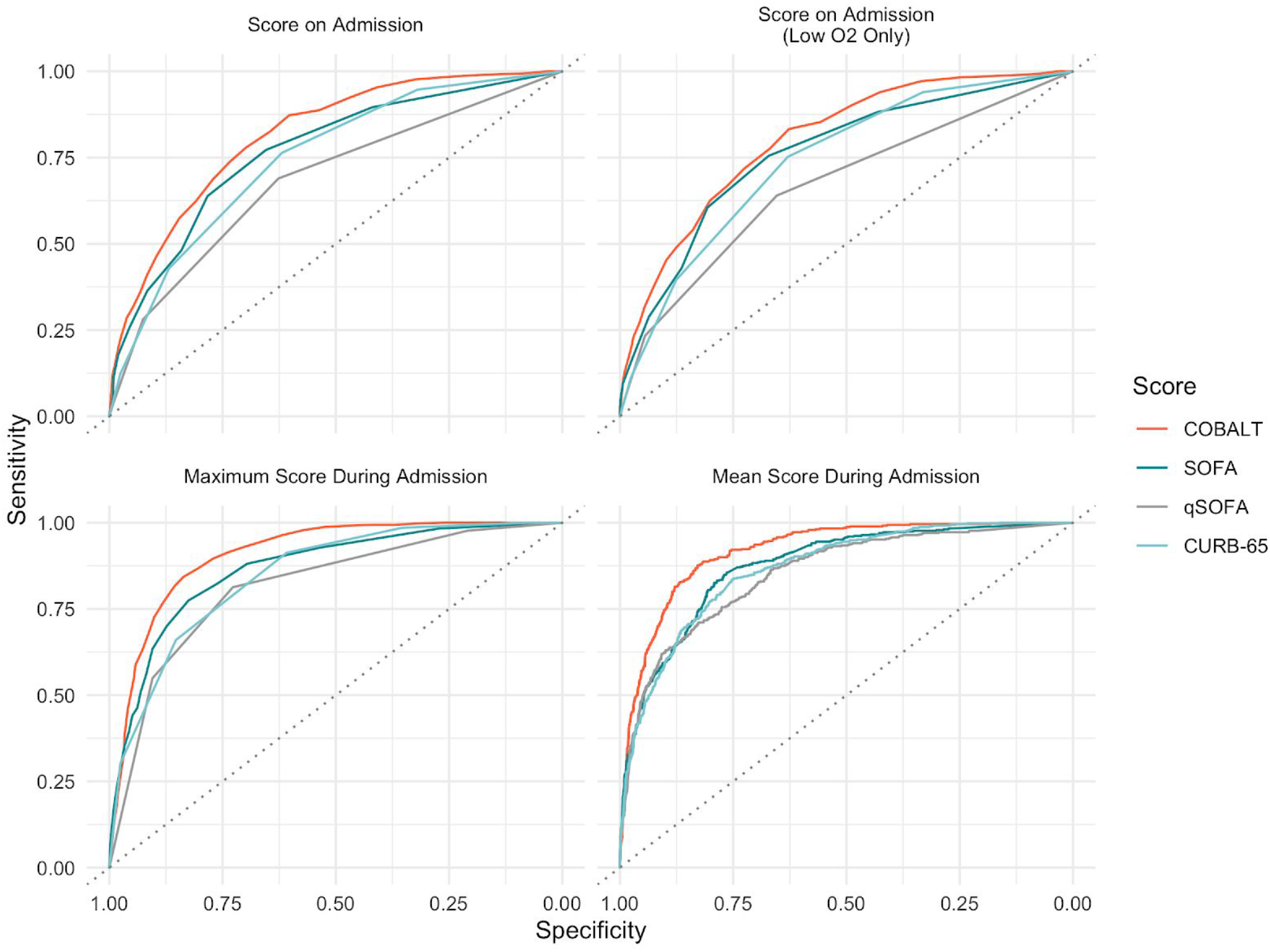
Comparison of AUC values for each mortality score for outcomes of death vs. other. The top two panels show the performance of scores on admission for (left) all patients and (right) those admitted on low-flow supplemental oxygen or room air only.

Because oxygen support level showed the strongest association with mortality, we confirmed that the COBALT score still performed well when only patients who were admitted on room air or low-flow oxygen were considered. The ROC curves for scores calculated at admission in this patient subset are shown separately in **Figure 2**. Performance declined slightly (AUCs: 0.80 for COBALT, 0.76 for SOFA, 0.74 for CURB-65, 0.67 for qSOFA) but the relative rankings of the four scores were consistent with those in the overall population.

**Figure 3:**
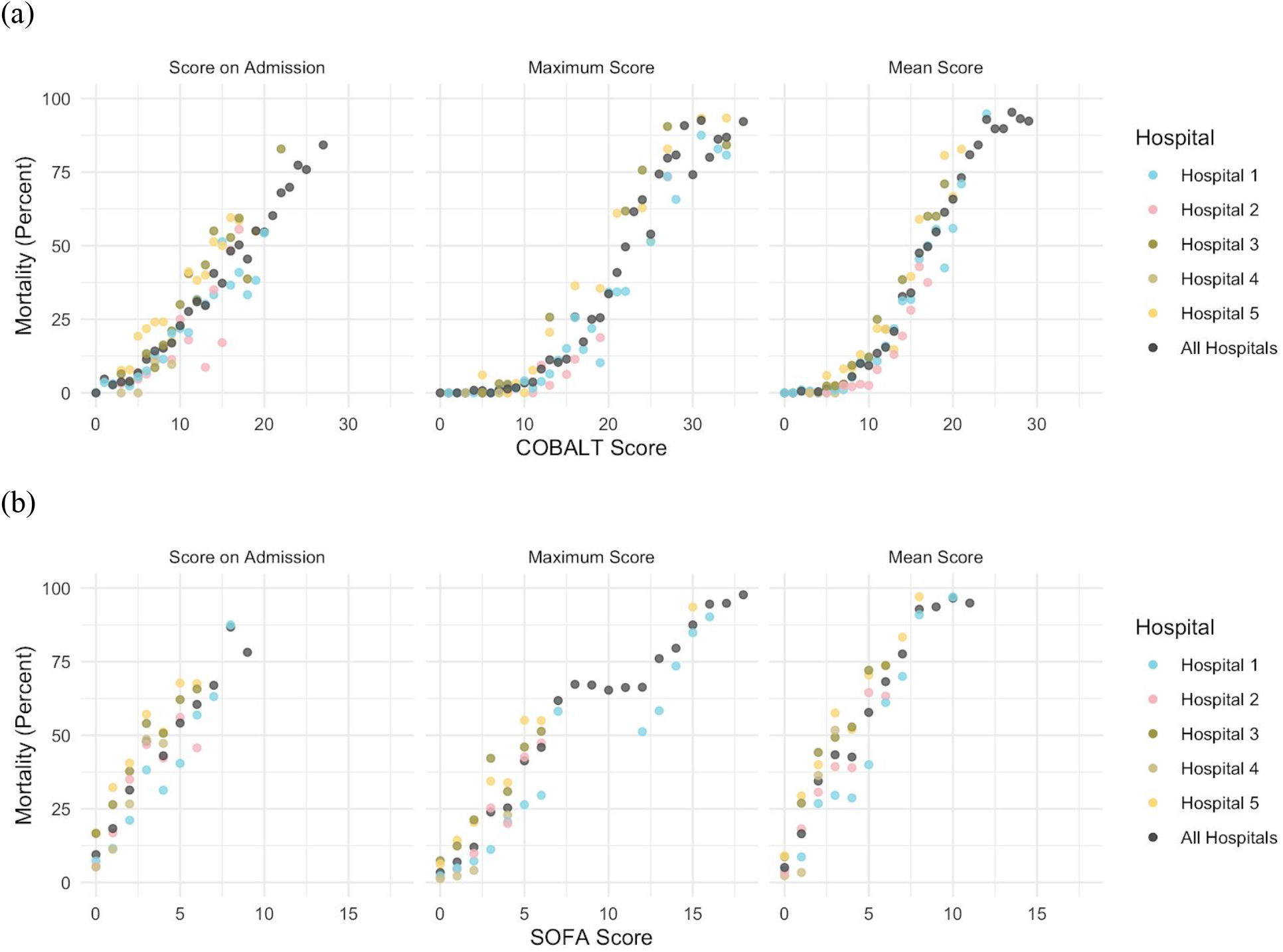
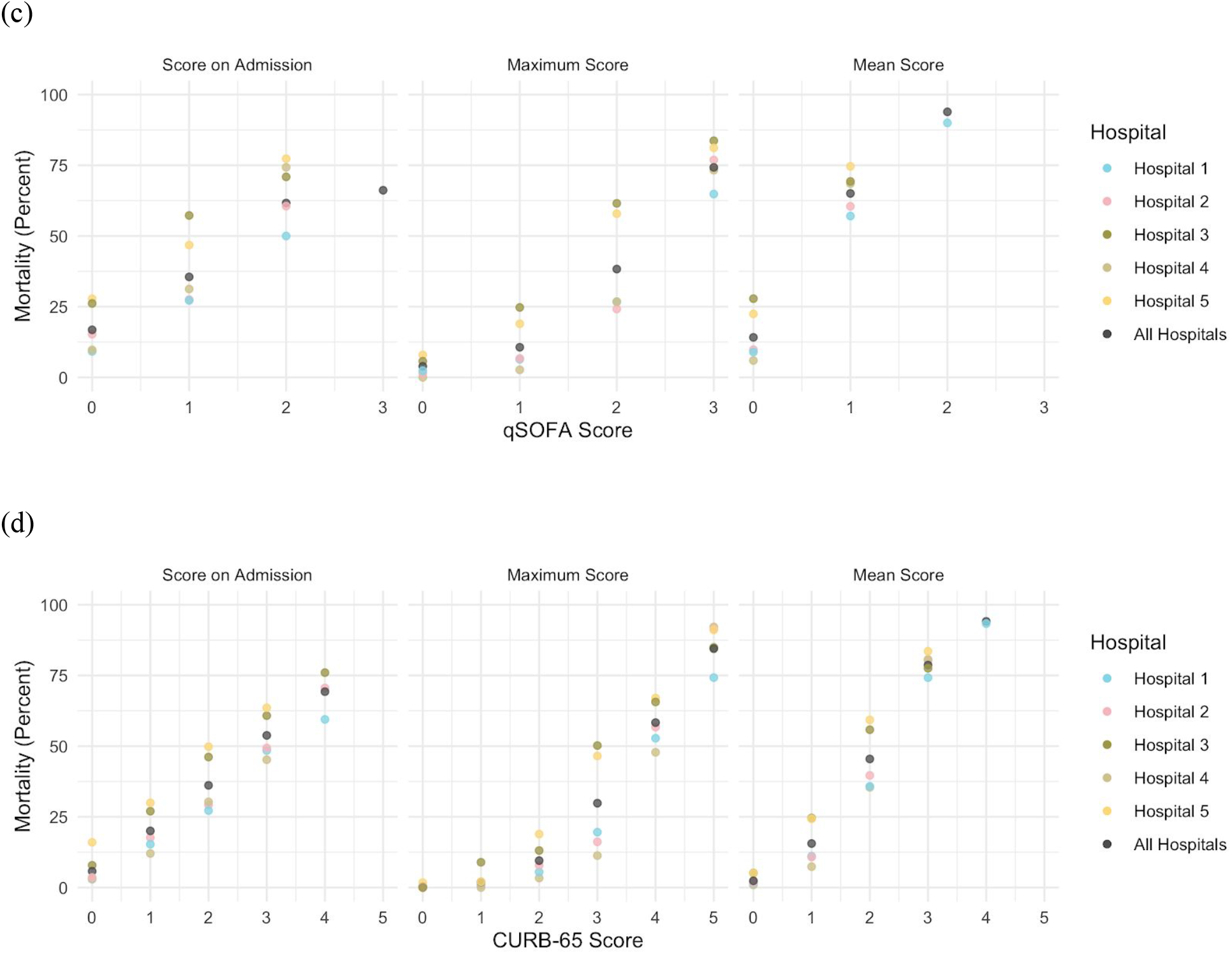
Mortality estimates in a COVID-19 population. Mortality estimates are shown at each possible score value for the (a) COBALT score (range: 0-36), (b) SOFA score (range: 0-24), (c) qSOFA score (range: 0-3), and (d) CURB-65 score (range: 0-5). Each colored dot represents the mortality at one score value at one of five hospitals. The dark gray dots represent the overall mortality at each score value in our entire patient population (n=4840). A dot is only present if sufficient patients (n>30) were present for that score value and hospital. Mean scores are usually non-integer values and are represented by their lower bounds (e.g. a mean score of 2.2 would be represented by a value of 2 in the graphs).

### Score-specific mortality calibration plots for COVID-19

Because even highly predictive scores can be poorly calibrated [29], we estimated mortality separately at each possible value of the COBALT, SOFA, qSOFA and CURB-65 scores (**Figure 3**). We also calibrated all four scores separately by hospital, as the five hospitals within our health system are located in different neighborhoods with different racial, ethnic, and socioeconomic compositions and have different baseline levels of COVID-19 mortality (**Figure S1**).

Mortality for the COBALT score is generally low (less than 5%) for maximum scores 0-10, moderate (5-25%) for maximum scores 11-18, high (25-75%) for maximum scores 19-24, and very high (greater than 75%) for maximum scores greater than 24. The tightest calibration (lowest variation in mortality among hospitals at each score level) occurred when the mean score was used. The COBALT score’s nearest competitor, SOFA, performs well in terms of AUC but suffers from high variation in mortality at low score ranges; there was no level of the SOFA score at which mortality was consistently below 5% in this population. The CURB-65 score, despite its lower AUC, is more tightly calibrated, particularly when used to assess mortality risk at the time of admission.

## Discussion

In this study, we present a novel clinical tool, the COBALT score, which estimates the risk of mortality in hospitalized COVID-19 patients with superior performance compared to three commonly-used mortality risk scores. To our knowledge, this is the first report to evaluate and calibrate widely-used mortality risk scores (SOFA, qSOFA, and CURB-65) in COVID-19. While the COBALT score outperforms these scores in terms of predictive ability, we recognize that some health providers would prefer to use scores with which they are already familiar. The calibration plots shown in **Figure 3** should assist providers who want to take this approach. While this calibration was internal to our health system, it provides a sense of the mortality risk and variation in mortality at each score level and ensures that uncertainty in mortality risk can be communicated simply and transparently, especially in pandemic settings.

Notably, the mortality cutoffs for SOFA, qSOFA and CURB-65 in this COVID-19 population differ substantially in some cases from those in earlier published mortality tables. For example, Ferreira et al report overall mortality of less than 10% for SOFA scores of 0-3 calculated at admission, whereas we found mortality rates as high as 50% in this population. Similarly, the original reference for qSOFA [5] estimated a 3-14 fold increase in mortality for patients with qSOFA scores of 2-3 relative to those with scores of 0-1. We found a more linear increase in mortality among different qSOFA score levels in this population, as well as considerable variation in mortality within each qSOFA score level. These findings reflect the importance of recalibrating mortality estimates for scores when they are used in contexts outside those in which they were originally validated.

Most of the variables strongly correlated with COVID-19 mortality, shown in Table 2, are consistent with the findings of prior studies. For example, we identified several laboratory values as potential risk score candidates, including lymphocyte percentage, D-dimer, troponin, lactic acid dehydrogenase (LDH), and blood urea nitrogen (BUN), all of which have been independently documented as prognostic factors in COVID-19 [11, 20-28]. However, several findings in Table 2 were unexpected, for example that Black race, Hispanic ethnicity, and BMI, which have been widely reported as predictive of COVID-19 prognosis, were only weakly associated with mortality [31]. In part, this was because we stratified the Gray tests by admission site; each of our five hospitals has a distinct racial and ethnic composition reflective of the surrounding neighborhood, and the effect of these variables within sites is likely weaker than that among them. Similarly, the effect of BMI was likely reduced because the majority of our admitted patients were overweight or obese (IQR 24.6-32.7), and our variable selection algorithm prioritized variables with distinct mortality patterns across quartiles. Still, it is interesting to note that variables that have been conclusively demonstrated to lead to increased COVID-19 hospitalization rates [32] do not necessarily lead to increased mortality once a patient is already in the hospital.

The fact that 36 out of the 48 variables we considered for our score showed significant individual associations with mortality raises the possibility that a multiplicity of mortality risk scores could potentially produce high predictive accuracy in COVID-19. Indeed, this is consistent with our observation that no two existing COVID-19 risk scores are exactly alike; the statistical algorithms deployed have chosen different variables in virtually all cases [12-16,30]. Our position is that, given this observation, less attention should be paid to improving predictive accuracy and more should be paid to a score’s practical aspects, such as calibration and ease of use. We have attempted to prioritize these concerns in our approach here.

Our study has several strengths. First, the Mount Sinai Health System admitted approximately 17% of all hospitalized COVID-19 patients in New York City during the first U.S. wave of the COVID-19 pandemic (March-April 2020). With five hospitals in three different boroughs represented in this study, we expect our results to be broadly representative of the clinical course of COVID-19 in hospital settings. Second, our data-driven approach to score design is novel and could serve as the basis for creation of simple mortality risk scores for other diseases. Third, the COBALT score can be calculated by hand at the bedside and uses only clinical observations and basic labs, a significant asset to triage and prognostication in resource-constrained settings. And finally, in addition to evaluating the score’s predictive performance, we also calibrated the score in our population of 4840 patients, both as one population as well as within each of five different sites of admission.

As a retrospective study within a single health system, our study also has some limitations. First, documentation of all kinds was inconsistent during the first wave of COVID-19, and the environments at different hospitals varied substantially. While it is unlikely that a laboratory result or medication administration was missed, inconsistencies in flowsheet documentation during this period could mean that the timings of different modes of oxygen administration were not always accurately captured. Second, the score may need to be calibrated separately when used outside hospital settings. For example, COBALT mortality thresholds reported here may differ when applied to skilled nursing facilities or to health systems in countries outside the U.S. Finally, we were unable to include two health system hospitals (Mount Sinai Beth Israel and Mount Sinai South Nassau) because, as of the time of this study, they did not yet use the Epic electronic health record system.

The COBALT score provides an easy-to-use, data-driven bedside tool to assess the risk of mortality in COVID-19 patients and outperforms other hospital mortality risk scores currently in widespread use. In addition, our analysis and recalibration of existing scores is, we believe, unique in the COVID-19 literature. We hope our score and the approach we took to design it will prove useful to our colleagues in the United States and throughout the world who continue to fight the COVID-19 pandemic.

## Data Availability

Any secondary use of study data is contingent upon approval by the Mount Sinai Data Use Committee. Please contact the corresponding author for further information.

## Acknowledgments

We thank Douglas Tremblay, Daniel Howell, Krishna Chokshi, and Leonard Naymagon, who manually validated intubation timings for the dataset used in this project. We thank Erick Scott for his careful attention to calibration in his own work and for providing relevant literature on the subject.Supplementary Material (Online)

---

Figure S1: Overall mortality and discharge outcomes by site of admission

Figure S2: Example mortality cumulative incidence plots for variables considered for final score

Figure S3: Example mortality cumulative incidence plots for variables not considered for final score

Table S1: Comparison of distributions of variables in datasets 1-3

